# A characteristic cerebellar biosignature for bipolar disorder, identified with fully automatic machine learning

**DOI:** 10.1101/2022.01.22.22269384

**Authors:** Georgios V. Thomaidis, Konstantinos Papadimitriou, Sotirios Michos, Evangelos Chartampilas, Ioannis Tsamardinos

## Abstract

**Backround:** Transcriptomic profile differences between patients with bipolar disorder and healthy controls can be identified using machine learning and can provide information about the potential role of the cerebellum in the pathogenesis of bipolar disorder.With this aim, user-friendly, fully automated machine learning algorithms can achieve extremely high classification scores and disease-related predictive biosignature identification, in short time frames and scaled down to small datasets.

**Method:** A fully automated machine learning platform, based on the most suitable algorithm selection and relevant set of hyper-parameter values, was applied on a preprocessed transcriptomics dataset, in order to produce a model for biosignature selection and to classify subjects into groups of patients and controls. The parent GEO datasets were originally produced from the cerebellar and parietal lobe tissue of deceased bipolar patients and healthy controls, using Affymetrix Human Gene 1.0 ST Array.

**Results:** Patients and controls were classified into two separate groups, with no close-to-the-boundary cases, and this classification was based on the cerebellar transcriptomic biosignature of 25 features (genes), with Area Under Curve 0.929 and Average Precision 0.955. Using 6 of the characteristic features (genes) discovered during the selection process, 99,6% of predictive performance was achieved. The 3 genes contributing most to the predictive power of the model (92,7% predictive performance) are also deregulated in temporal lobe epilepsy. KEGG analysis revealed participation of 4 identified features in 6 pathways which have been associated with bipolar disorder.

**Conclusion:** 93% Area Under Curve, 96% Average Precision, and complete separation between unaffected controls and patients with bipolar disorder, were achieved in ∼2 hours. The cerebellar transcriptomic biosignature suggests a potential genetic overlap with temporal lobe epilepsy and new genetic contributions to the pathogenesis of bipolar disorder.

## 1. Background

Bipolar disorder (BD) is a mood disorder characterized by unusual fluctuations of mood, thinking, activity and sleep patterns, classified in six subtypes [1] (with bipolar disorder types 1 and 2 the most prevalent) and presented as a constellation of phenotypes, with a variety of cognitive and behavioral features [2]. It is a highly hereditary disease, running in families, with an early onset, unpredictable course and detrimental impact due to the great risk of fatal self-destructive events, long term disability and great financial and social burden, despite existing pharmacological and psychotherapeutic treatment strategies [3]. For these reasons, the neuroanatomy [4], neurogenetics and neurobiology [5] of BD are fields of intense research concerning all brain areas and of paramount importance for 45 million patients globally [6]. In this context, the cerebellum is a relatively recent target of neurogenetics research in BD, with its main functional roles related to modulation of movement, to emotion and to cognition [7]. Research has linked the cerebellum to emotional, cognitive and affective processing and their disruption in mood disorders [7], [8]. Structural [9], [10], [11], [12], [13], [14], [15], functional [7], [8], [14], [16], [17], [18], [19], neurotransmission [20], [21], metabolic [22], [23], [24], [25] and transcriptomic [26], [27], [28], [29] alterations in the cerebellum in BD point to the cerebellum’s particular role in the affected brain-wide networks.

Machine learning is now gradually being used in psychiatry, in order to optimize genetic analysis [30], [31], to highlight the most characteristic differences among groups of patients and controls, and to confirm their importance for diagnostic classification into these groups. These complex classification algorithms, produce genetic signatures using data from the analysis of samples from living tissue, blood, saliva, as well as from postmortem brain tissue (prefrontal cortex) [31]. The data include SNP (5 studies) [30, table 1.] and transcriptomics (2 studies) [31], [32] analysis results. In this context, transcriptomic data analysis can contribute greatly to psychiatric research [33]. Data from the less explored area of the cerebellum can add new and important biosignatures to the puzzle of BD pathogenesis and progression, and potentially to treatment response and resistance. The current study is, as far as we know, the first where autoML and transcriptomic data from the cerebellum were used for biosignature identification and patient classification.

## 2. Aims of the study

The aim of this analysisis the selection of characteristic transcriptomic biosignatures of bipolar disorder in the cerebellum, using the autoML platform for optimal performance. The features identified could facilitate the discovery of the genetic networks related to BD, highlight their importance at the local and brain-wide network levels and explore a potential genetic overlap with other central nervous system (CNS) disorders.

## 3. Methods

For this study, we applied the fully automatic machine learning (autoML) platform Just Add Data Bio (JADBIO) [34] on public transcriptomic data from previous studies [30], [31] that analyzed the transcriptomic profiles of the cerebellum and parietal cortex of postmortem brain tissues and produced a set of biosignatures [29]. Patient and control groups were homogenized by tissue sample location (cerebellum), psychiatric diagnosis, sex and age. The autoML system has a simple, user-friendly interface and has been created for direct application on low-sample, high-dimensional databases. The issue of reliability of sophisticated, non-linear machine learning analyses on small sample data (∼30), has been specifically and thoroughly addressed theoretically [35] and for applications on omics datasets in precision oncology [34].

The platform is automatically trained and evaluated (tested), in order to identify highly optimized predictive and classification models, using characteristic biosignature profiles. When provided with a well-defined set of features (e.g., data from transcriptomic, biochemical, neuroimaging, psychometric or symptom intensity studies), it can produce a very small subset of predictors (biosignatures / characteristic features) selected from among these, leading to increased predictive power performance. In studies which work with binary classification (e.g., the current study, in which classification into the two groups of BD patients and controls took place using transcriptomic data), the classification boundary is defined by the most statistically significant combination of biosignatures (the characteristic biosignature), which distinguishes patients from controls. System applicability has been tested for diagnostic classification and time to event prediction, producing robust classification, biosignature identification and prediction results (AUCs 85-95%), using data from oncology, neurology and psychiatry [36-40].

### 3.1 Data acquisition

Publicly available data have been used, from the online BioDataome database [41], which is constructed by uniformly preprocessed, disease-annotated omics data from GEO and RECOUNT databases, based on a uniform preprocessing pipeline, described in detail at the BioDataome documentation page [42]. We analyzed the BioDataome csv. which corresponds to the GEO dataset GSE35978, a. containing expression data from the human cerebellum (produced from GSE35974) and parietal cortex, b. from post mortem brain tissue samples, c. extracted from unaffected subjects and schizophrenic, bipolar and depressed patients, d. from the Stanley Foundation Brain Collection [43]. The expression data were obtained by microarray analysis using the “[HuGene-1_0-st] Affymetrix Human Gene 1.0 ST Array [transcript (gene) version]”. The dataset was initially used for the analyses by Chen C et al [27], [28]. Technical details about the initial postmortem sample (age, Ph, postmortem interval, sex etc) are available at the EMBL-EBI page for E-GEOD-35978 [44], [45]. Demographic data about a. race/ethnicity, b. side of brain of the samples, c. Bipolar Disorder types, d. Occurrence of psychotic features and e. cause of death of the participants, are available at the Array Collection description [46]. Inclusion criteria and diagnostic methodology for the samples of the Stanley Foundation Brain Collection, are described at the Tissue Repository information page [47].

### 3.2 Data Processing

#### 3.2.1 Dataset selection and homogenization

Data have been downloaded in .csv format from the BioDataome database. The preprocessed file includes data for 144 samples from the cerebellum and 168 samples from the parietal cortex. The 144 cerebellum samples include unaffected subjects and patients with bipolar disorder, schizophrenia and depression (SI, information on GSE35974 and GSE35978). From the cerebellum group, all 50 unaffected subjects and 37 bipolar disorder patients (sex: females/ males, age span: 20 - 70) were initially chosen (SI, Images 1A-1B and 1C-1D). From the initial heterogeneous groups of patients and controls, a number of subjects were removed, and two new, smaller groups of affected / unaffected subjects were produced, matched for sex (female / male) and for age. At the same time, we aimed to exceed (as much as the sample sizes allowed) the minimum threshold of 30 subjects per group required for the machine learning analysis (SI, Images 2A-2B and 2C-2D). The final dataset includes the following two groups: Group A with 35 bipolar patients (18 female and 17 male) and Group B with 37 unaffected controls (19 female and 18 male). The small size of available data excluded the possibility of testing after the initial training; this was balanced by the extremely high AUCs produced during the initial (training) analysis. During the initial microarray analysis, a number of transcriptomes were used as controls [27], [28]. These have been identified and removed from the csv. of the analysis, and the final datasheet (Diagnosed Subjects x Features) consequently produced. The datasets are 2D matrices (features/ genes x diagnosis for any given subject, unaffected or patient).

#### 3.2.2 Feature selection and biosignature construction

For the analysis, data were uploaded to JADBIO version 1.4.14 (April 2021) and the binary classification (categorical) functionality of the platform was employed. The classification process is based on the Statistically Equivalent biosignatures (SES) method, with Support Vector Machines, Random Forest, and Penalized Linear Models algorithms. [34], [39]. For the given 2D matrices, the predicted outcome is diagnosis (Bipolar or Unaffected), and the metric chosen for optimization is the AUC. Preprocessing used Constant Removal Standardization.

Feature selection was performed using LASSO (Least Absolute Shrinkage and Selection Operator) Feature Selection (penalty=0.0, lambda=5.509e-02). The analysis protocol followed has been a repeated 10-fold cross validation without dropping (max. repeats = 20), with 596 configurations, 5760 predictive models trained and 83440 predictive models omitted (total 89200). The chosen predictive algorithm uses Ridge Logistic Regression (with penalty hyper-parameter lambda = 10.0). The overall process applies the Bootstrap Bias Corrected Cross Validation, a protocol for algorithm hyper-parameter tuning during performance estimation and multiple tie adjustment [34], [39]. Time to complete was 2 hours 16 minutes. The technical analysis report is in SI-Appendix-1.

## 4. Results

### 4.1. Classification between BD patients and unaffected controls

The AutoML classification analysis produced a Ridge Logistic Regression model with high AUC for the positive class bipolar (93%), based on 25 characteristic biosignatures. AUC, and Average Precision (AP) values and Confidence Intervals(CIs), Receiver Operating Characteristic Curve (ROC curve) and main optimized classification threshold dependent metrics for Accuracy / Balanced Accuracy are shown in Image 1.The BD and HC groups produced are completely separate and coherent in the Uniform Manifold Approximation and Projection (UMAP) plot (Image 2).

**Image 1.**
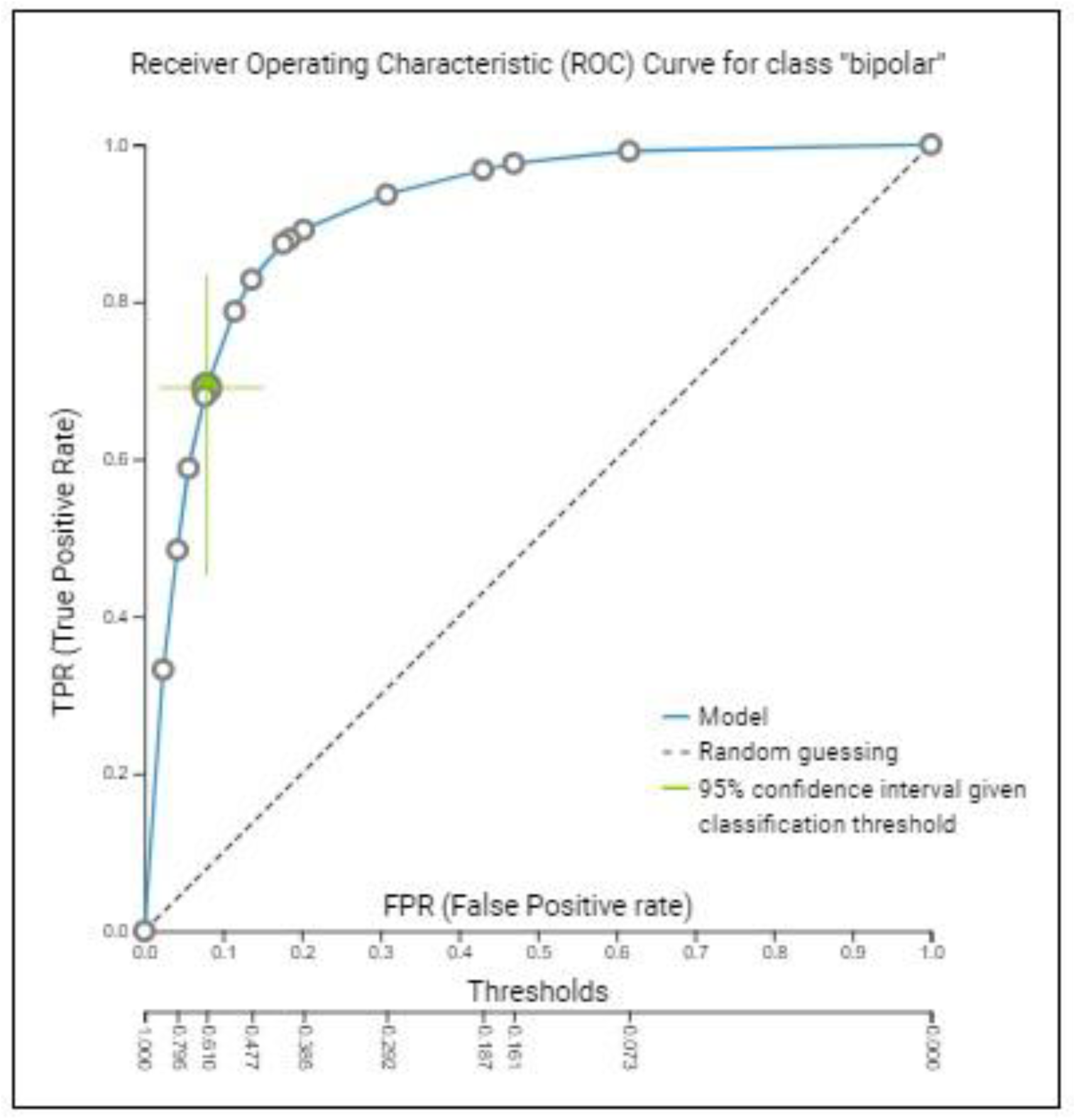
Using the best performing model option in the platform, the AUC for the positive class bipolar is 0.929 (∼93%), with a 95% CI between 0.868 - 0.977, and the AP is 0.955, with a 95% CI between 0.914 - 0.986. Accuracy has been calculated at 0.843, Precision at 0.906 and Specificity at 0.921 (full data in SI, Image 3). The classification threshold (0,61) has been optimized and determined for Accuracy / Balanced Accuracy. Classification as positive is performed when out-of-sample predicted probability is above this given threshold (0,61).

**Image 2.**
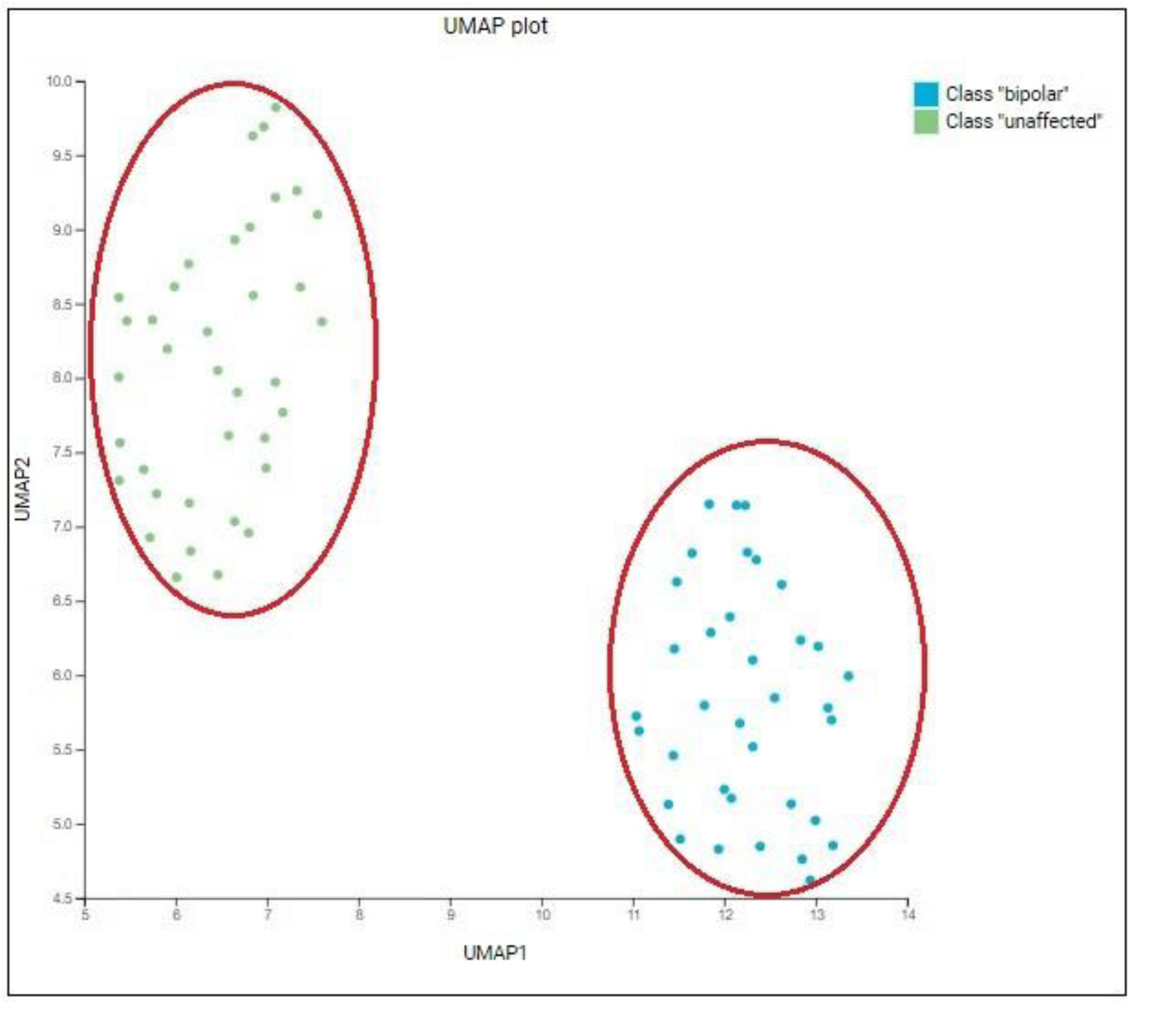
Complete separation of BD patients from unaffected controls, in UMAP plots based on all the25 selected biosignatures. In the Box Plot contrasting the cross-validated predicted probability of belonging to a specific class against the actual class of the samples, the medians are ∼0,72 for the class “bipolar” and ∼0,18 for the class “unaffected” (SI, Image 4).

### 4.2 Biosignature identification

The algorithm selected the most important 25 out of the 28869 features of the dataset. These 25 features (characteristic features) constitute the reference signature, used for the classification between BD and controls. Inclusion of the 6 most important features (gene transcriptomes from **RNU6-576P, MIR194-2, GDPD5, CARD16, RABGGTA, KREMEN2**) achieves predictive performance (PP) 99,603%. Inclusion of the most important feature RNU6-576P leads to 76,6% PP, inclusion of the first and second (MIR194-2) most important feature achieves 85,8% PP, and additional inclusion of the third most important feature (GDPD5) achieves 92,7% PP. The progressive feature inclusion plot for the 6 most important out of the 25 identified features is presented in Image 3.

**Image 3.**
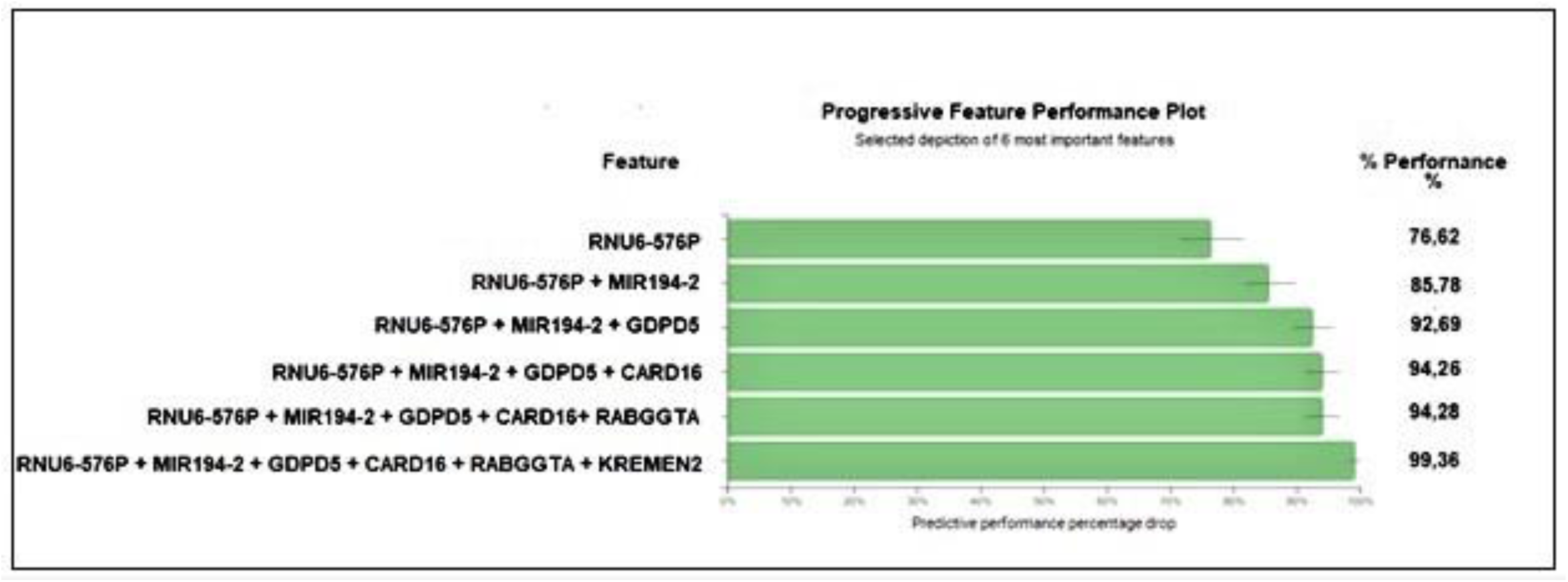
Progressive feature inclusion plot. This plot reports the predictive performance (in percentage) that can be achieved by using only the 6 of the 25 characteristic features (mentioned above). The features are added one at a time, starting from the most important and ending with the complete signature. Grey lines indicate 95% confidence intervals. In this image the predictive performance of the 6 most important features is presented.

### 4.3 KEGG analysis

We applied KEGG analytical tools to the characteristic features with KEGG identifiers. Four of these genes (**MIR194-2, CARD16, ASIC3, H4C1**) are involved in 7 KEGG pathways: MIR194-2 in the hsa05206 pathway (“MicroRNAs in cancer”) [48], CARD16 in the hsa04621 pathway (“NOD-like receptor signaling”) [49], ASIC3 in the hsa04750 pathway (“Inflammatory mediator regulation of TRP channel”) [50] and H4C1 in 4 pathways: hsa05034 (“Alcoholism”) [51], hsa04613 (“Neutrophil extracellular trap formation”) [52], hsa05203 (“Viral carcinogenesis”) [53], hsa05322 (“Systemic lupus erythematosus - Homo sapiens”) [54].

## 5. Discussion

### 5.1 Main Findings

#### 5.1.1 Genetic Biosignature genes and Neuropsychiatric Disease

The classification between the Bipolar and unaffected control groups was completed in <1 hour, with accuracy ∼93% and without overlaps between the produced sets of individuals. The Welsh t-test for the 6 most important genes established that the differences in expression between patients with bipolar disorder and unaffected controls are statistically meaningful (SI, Image 5). Classification using the JADBIO platform can be considered a reliable means and produces robust results, with potential research interest. The single most important identifier was by far the RNU6-576P small non-coding RNA, accounting for ∼77% of total feature importance. The three most important features The three most important features according to their performance are RNU-576P, MIR194-2, and GDPD5. There are at least 21 known or probable functional roles of RNU6-576P, MIR194-2, and GDPD5 in the nervous system and in CNS diseases (including the gene aliases from Gene Cards). 36 related articles are presented in SI-Appendix 1.

The most consistent and important finding is that RNU6-576P, MIR194-2 and GDPD5 have been associated with epilepsy (though not until now with bipolar disorder). Both epilepsy and bipolar disorder are characterized by episodic functional deregulation in the CNS [55], co-occur [56], share common symptoms and precipitating factors [57], [58], their treatment with antiepileptics / mood stabilizers partially overlaps [59], and potential pathophysiological links have been proposed recently [60] regarding aberrant neuronal excitation-inhibition related to ANK-3 gene expression. Epileptiform EEG discharges are connected to progress and worse course of disease in BDII patients [61] and manic symptoms are more common in patients with temporal lobe epilepsy [57]. ANK3 belongs to a cluster of genes with altered expression patterns in the cerebellar vermis, in patients with bipolar disorder [26], [62]. Significantly, alterations in RNU6-576P and MIR194-2 expression are connected to temporal lobe epilepsy [63], [64], [65], [66], [67], which shares the most common symptoms and pathways with Bipolar Disorders I and II [56] - [61].

The role of small non-coding RNAs and pseudogenes is a new area of intense research in relation to the onset of psychotic disorders, depression and bipolar disorder [64], [65] and their participation in the epigenetic modification of DNA [48]. RNU6-576P is the most overexpressed small non-coding mRNA in the hippocampus of patients with mesial temporal lobe epilepsy [63] and the most important identifying biosignature in the cerebellum of BD patients in this study. The role of MIR-194-2 expression in epilepsy has been studied in the greatest detail, and a constant pattern of down-regulation has been documented, in various epilepsy studies [66] - [69].

#### 5.1.2. KEGG Analysis results and Neuropsychiatric Disease

Associations of the identified KEGG pathways with neuropsychiatric diseases or proposed pathophysiological or treatment mechanisms, have been found in several studies and the results are presented analytically.

a) **hsa05206 pathway includes** MIR194-2 gene and also miR-34a gene. miR-34a expression alterations in the cerebellum have been connected to bipolar disorder in previous studies in the same post-mortem sample [29], [70]. Secondarily, the hsa05206 pathway, has been connected to the mechanism of action of saikogenin G, a bioactive ingredient of the traditional antidepressant treatment Radix Bupleuri in Chinese medicine [71], but has not been connected to response to lithium therapy in bipolar disorder [72].
b) **hsa04621 pathway** (includes CARD16 gene) implicates immunological deregulation in bipolar disorder [73], schizophrenia [74], [75] depression [76] and epilepsy [77] and is a target pathway for certain antiepileptics [78].
c) **hsa04750 pathway** (includes ASIC3 gene), has been connected to schizophrenia [79], post-stroke depression [80] and to metabolic syndrome in bipolar disorder and schizophrenia [81].
d –g) (pathways involving H4C1)

**hsa05322**, **hsa05034**, **hsa05203** and **hsa04613 pathways**, have been all associated with a genetic risk for depression [82], with many mechanisms mediated by immunological processes [83].

Also, **hsa04613** has been separately connected [84] to depression and to fatigue (in patients receiving chemotherapy for cancer) [85]; fatigue is also a characteristic symptom of depression. **hsa05203** has been separately connected to Post Traumatic stress Disorder, a major predictor for depression [86], epigenetic contributions to human behavior [87], risk for first episode psychosis [88] and schizophrenia [89] and **hsa05322** has been separately associated with oxidative stress and cognitive function in schizophrenia [90] and antipsychotics-induced parkinsonism [91].

Concisely, 6 out of 7 identified KEGG pathways, are associated with bipolar disorder and major psychiatric diseases (depression, psychosis) -which share common phenotypical features with bipolar disorder- and with epilepsy. The **hsa05206 pathway** is of particular interest, as it includes also miR-34a gene; Both ANK3 (Ankyrin-3) and CACNB3 (voltage-dependent L-type calcium channel subunit beta-3) genes, are directly targeted by miR-34a [29], [70]. ANK3 and could be connected to the neurobiology of bipolar disorder [70], [92], [93], [94], [95].

### 5.2 Limitations of the study

The present study was based on a relatively small sample of patients with BD Types Ι and II, with an increased analogy of deaths from suicide, and was based on post-mortem tissue sampling and microarray analysis. Genetic differences between patients with BD I and BD II have been suggested [96], [97], [98], [99], [100], using family databases, but neuroimaging differences have not been confirmed [101]. The bipolar spectrum is highly heterogeneous, with many different biotypes and their probable neurobiological causes [4], [102], [103], [104], [105]; different biotypes can be fully represented only in large samples [4]. The BD group of 37 subjects includes 7 patients who had committed suicide, a number close to known prevalence of death by suicide in BD. Suicide mainly occurs during the depressive state of the disease and – occasionally – during a manic episode, and could be connected to certain patterns of gene expression [104] and biosignature differences [106], found also in the cerebellum. Also. its frequency can vary depending on the depressive, manic or mixed state of the disease [107]. Finally, future studies, based on progresses in knowledge of the cerebellar transcriptomic landscape [108], using microarray [109], RNA-seq and other methods, could provide additional insights in the neurobiology of BD in the cerebellum and the region specificity of our and future findings. Further study is very important, as the genetic characteristics of post-mortem brain tissue samples could be extremely complex in the same area [110] and divergent from the same characteristics of the living brain, in health and disease; still they remain one of the cornerstones of research on the neurobiology of the CNS and its disorders [111], [112], [113], [114].

## Supporting information

https://drive.google.com/file/d/17IRmrroNB-CM2H2WKrhQtQPaC3jQbgia/view?usp=sharing

## Data Availability

All data produced in the present study are available upon reasonable request to the authors

https://docs.google.com/document/d/13ehWWGhczwTii47jcTaGUwdJTu8aB3ZjT1D89_mpY3I/edit?usp=sharing

## Abbreviation List

AutoML: Automatic Machine Learning
GEO: Gene Expression Omnibus
AUC: Area Under Curve
AP: Average Precision
TLE: Temporal Lobe Epilepsy
BD: Bipolar Disorder
CNS: Central Nervous System
JADBIO: Just Add Data Bio
SES: Statistically Equivalent bioSignatures
LASSO: Least Absolute Shrinkage and Selection Operator
ROC: Receiver Operating Characteristic (curve)
CI: Confidence Interval
UMAP: Uniform Manifold Approximation and Projection

## Declarations

### Authors’ contributions

G.V.T. designed the experiment, interpretated the results and wrote the main manuscript text. I.T. designed the software for the experiment and reviewed the manuscript. S.M., K.P. and E.C. performed equally the acquisition and analysis of data. S.M and K.P contributed equally to the methods section.

## Authors’ information

Ioannis Tsamardinos is Professor of Data Science and Bioinformatics in the University of Crete, Department of Computer Science, Heraclion, Greece. He has more than 10.000 citations in the field.

Georgios V. Thomaidis is a Psychiatrist (MD, MSc), researcher and trainer of psychiatric residents, with post-graduate studies (MSc level) in Nanoscience and Complex Systems. He practices Psychiatry at the Department of Psychiatry of Katerini General Hospital (ex. Psychiatric Hospital of Petra Olympus, Pieria). Sotirios Michos, PhD Electronic Engineering, is a researcher in the fields of Network Science and Bioinformatics.

Konstantinos Papadimitriou, is a molecular biologist (MSc) and resident Psychiatrist in the Psychiatric Hospital of Thessaloniki, Greece.

Evangelos Chartampilas is a Clinical Radiologist, researcher and trainer of Radiology residents at the Radiology Department Aristotle University of Thessaloniki, Greece.

## Preprint

A preprint version of the article can be found at: https://www.medrxiv.org/content/10.1101/2022.01.22.22269384v1

