## Supplementary material for "A characteristic cerebellar biosignature for bipolar disorder, identified with fully automatic machine learning": https://drive.google.com/file/d/17IRmrroNB-CM2H2WKrhQtQPaC3jQbgia/view?usp=sharing

1. Greek National Health System, Psychiatric Department, Katerini General Hospital, Katerini, Greece, 2. Greek National Health System, G. Papanikolaou General Hospital, Organizational Unit - Psychiatric Hospital of Thessaloniki, Thessaloniki, Greece, 3. Department of Computer Science, University of Crete, Heraclion, Greece, 4. Laboratory of Radiology, AHEPA General Hospital, University of Thessaloniki, Thessaloniki, Greece

***Corresponding author**

#### Abstract

**Backround:** Transcriptomic profile differences between patients with bipolar disorder and healthy controls can be identified using machine learning and can provide information about the potential role of the cerebellum in the pathogenesis of bipolar disorder.With this aim, user-friendly, fully automated machine learning algorithms can achieve extremely high classification scores and disease-related predictive biosignature identification, in short time frames and scaled down to small datasets.

**Method:** A fully automated machine learning platform, based on the most suitable algorithm selection and relevant set of hyper-parameter values, was applied on a preprocessed transcriptomics dataset, in order to produce a model for biosignature selection and to classify subjects into groups of patients and controls. The parent GEO datasets were originally produced from the cerebellar and parietal lobe tissue of deceased bipolar patients and healthy controls, using Affymetrix Human Gene 1.0 ST Array.

**Results:** Patients and controls were classified into two separate groups, with no close-to-the-boundary cases, and this classification was based on the cerebellar transcriptomic biosignature of 25 features (genes), with Area Under Curve 0.929 and Average Precision 0.955. Using 6 of the characteristic features (genes) discovered during the selection process, 99,6% of predictive performance was achieved. The 3 genes contributing most to the predictive power of the model (92,7% predictive performance) are also deregulated in temporal lobe epilepsy. KEGG analysis revealed participation of 4 identified features in 6 pathways which have been associated with bipolar disorder.

**Conclusion:** 93% Area Under Curve, 96% Average Precision, and complete separation between unaffected controls and patients with bipolar disorder, were achieved in ~2 hours. The cerebellar transcriptomic biosignature suggests a potential genetic overlap with temporal lobe epilepsy and new genetic contributions to the pathogenesis of bipolar disorder.

**Keywords**
Bipolar, Disorder, Temporal, Lobe, Epilepsy, RNU6-576P, AutoML, Machine, Learning, Psychiatry

A. Information about the GSE35974 and GSE35978
and BioDataome datasets and studies produced.

Original Dataset :

Expression data from the human cerebellum and parietal cortex brain.

Gene Expression Omnibus Accession Viewer link for GSE35974
https://www.ncbi.nlm.nih.gov/geo/query/acc.cgi?acc=GSE35974
Gene Expression omnibus Accession Viewer link for GSE35978:
https://www.ncbi.nlm.nih.gov/geo/query/acc.cgi?acc=GSE35978

Protocol Information for GSE35974
https://www.ebi.ac.uk/arrayexpress/experiments/E-GEOD-35974/protocols/
Protocol Information for GSE35978
https://www.ebi.ac.uk/arrayexpress/experiments/E-GEOD-35978/protocols/

**Full Subject list for GSE35978**
https://www.ebi.ac.uk/arrayexpress/experiments/E-GEOD-35978/samples/?s_page=1&s_pagesize=500&s_sortby=col_12&s_sortorder=ascending

BioDataome Processed Dataset
http://dataome.mensxmachina.org/data/Homo%20sapiens/GPL6244/GSE35978.csv

BioDataome Process Documentation:
http://dataome.mensxmachina.org/docs

| 1A - Unaffected (male) | | | | |
|---|---|---|---|---|
| **No** | **Sample ID** | **Sex** | **Age** | **Diagnosis** |
| 1 | GSM878305 1 | male | 30 | unaffected |
| 2 | GSM878293 1 | male | 30 | unaffected |
| 3 | GSM878275 1 | male | 30 | unaffected |
| 4 | GSM878257 1 | male | 35 | unaffected |
| 5 | GSM878252 1 | male | 35 | unaffected |
| 6 | GSM878239 1 | male | 35 | unaffected |
| 7 | GSM878337 1 | male | 40 | unaffected |
| 8 | GSM878300 1 | male | 40 | unaffected |
| 9 | GSM878261 1 | male | 40 | unaffected |
| 10 | GSM878295 1 | male | 45 | unaffected |
| 11 | GSM878294 1 | male | 45 | unaffected |
| 12 | GSM878268 1 | male | 45 | unaffected |
| 13 | GSM878259 1 | male | 45 | unaffected |
| 14 | GSM878258 1 | male | 45 | unaffected |
| 15 | GSM878221 1 | male | 45 | unaffected |
| 16 | GSM878321 1 | male | 50 | unaffected |
| 17 | GSM878310 1 | male | 50 | unaffected |
| 18 | GSM878309 1 | male | 50 | unaffected |
| 19 | GSM878306 1 | male | 50 | unaffected |
| 20 | GSM878301 1 | male | 50 | unaffected |
| 21 | GSM878290 1 | male | 50 | unaffected |
| 22 | GSM878288 1 | male | 50 | unaffected |
| 23 | GSM878273 1 | male | 50 | unaffected |
| 24 | GSM878240 1 | male | 50 | unaffected |
| 25 | GSM878230 1 | male | 50 | unaffected |
| 26 | GSM878338 1 | male | 55 | unaffected |
| 27 | GSM878280 1 | male | 55 | unaffected |
| 28 | GSM878242 1 | male | 55 | unaffected |
| 29 | GSM878236 1 | male | 55 | unaffected |
| 30 | GSM878336 1 | male | 60 | unaffected |
| 31 | GSM878322 1 | male | 60 | unaffected |

| 1B - Unaffected (female) | | | | |
|---|---|---|---|---|
| **No** | **Sample ID** | **Sex** | **Age** | **Diagnosis** |
| 1 | GSM878354 1 | female | 30 | unaffected |
| 2 | GSM878353 1 | female | 30 | unaffected |
| 3 | GSM878314 1 | female | 35 | unaffected |
| 4 | GSM878313 1 | female | 35 | unaffected |
| 5 | GSM878286 1 | female | 35 | unaffected |
| 6 | GSM878260 1 | female | 35 | unaffected |
| 7 | GSM878307 1 | female | 40 | unaffected |
| 8 | GSM878245 1 | female | 40 | unaffected |
| 9 | GSM878347 1 | female | 45 | unaffected |
| 10 | GSM878263 1 | female | 45 | unaffected |
| 11 | GSM878226 1 | female | 45 | unaffected |
| 12 | GSM878281 1 | female | 50 | unaffected |
| 13 | GSM878335 1 | female | 55 | unaffected |
| 14 | GSM878334 1 | female | 55 | unaffected |
| 15 | GSM878333 1 | female | 55 | unaffected |
| 16 | GSM878332 1 | female | 55 | unaffected |
| 17 | GSM878331 1 | female | 55 | unaffected |
| 18 | GSM878330 1 | female | 55 | unaffected |
| 19 | GSM878345 1 | female | 70 | unaffected |

Images 1A – 1B. Preliminary GSE35978 dataset with 50 unaffected subjects.

| 1 – C Bipolar (male) | | | | |
|---|---|---|---|---|
| No | Sample ID | Sex | Age | Diagnosis |
| 1 | GSM878222 1 | male | 20 | bipolar |
| 2 | GSM878348 1 | male | 30 | bipolar |
| 3 | GSM878311 1 | male | 30 | bipolar |
| 4 | GSM878231 1 | male | 30 | bipolar |
| 5 | GSM878214 1 | male | 30 | bipolar |
| 6 | GSM878356 1 | male | 35 | bipolar |
| 7 | GSM878272 1 | male | 35 | bipolar |
| 8 | GSM878229 1 | male | 35 | bipolar |
| 9 | GSM878227 1 | male | 40 | bipolar |
| 10 | GSM878289 1 | male | 45 | bipolar |
| 11 | GSM878248 1 | male | 45 | bipolar |
| 12 | GSM878329 1 | male | 50 | bipolar |
| 13 | GSM878326 1 | male | 50 | bipolar |
| 14 | GSM878264 1 | male | 50 | bipolar |
| 15 | GSM878220 1 | male | 50 | bipolar |
| 16 | GSM878342 1 | male | 55 | bipolar |
| 17 | GSM878233 1 | male | 55 | bipolar |
| 18 | GSM878255 1 | male | 60 | bipolar |
| 19 | GSM878251 1 | male | 65 | bipolar |

| 1 – D Bipolar (female) | | | | |
|---|---|---|---|---|
| No | Sample ID | Sex | Age | Diagnosis |
| 1 | GSM878344 1 | female | 25 | bipolar |
| 2 | GSM878267 1 | female | 30 | bipolar |
| 3 | GSM878284 1 | female | 35 | bipolar |
| 4 | GSM878235 1 | female | 35 | bipolar |
| 5 | GSM878243 1 | female | 40 | bipolar |
| 6 | GSM878298 1 | female | 45 | bipolar |
| 7 | GSM878271 1 | female | 45 | bipolar |
| 8 | GSM878269 1 | female | 45 | bipolar |
| 9 | GSM878339 1 | female | 50 | bipolar |
| 10 | GSM878282 1 | female | 50 | bipolar |
| 11 | GSM878278 1 | female | 50 | bipolar |
| 12 | GSM878225 1 | female | 50 | bipolar |
| 13 | GSM878270 1 | female | 55 | bipolar |
| 14 | GSM878325 1 | female | 60 | bipolar |
| 15 | GSM878244 1 | female | 60 | bipolar |
| 16 | GSM878232 1 | female | 60 | bipolar |
| 17 | GSM878266 1 | female | 65 | bipolar |
| 18 | GSM878265 1 | female | 65 | bipolar |

Images 1C – 1D. Preliminary GSE35978 dataset with 37 bipolar disorder patients.

| 2A - Unaffected (male) | | | | |
|---|---|---|---|---|
| No | Sample ID | Sex | Age | Diagnosis |
| 1 | GSM878305 1 | male | 30 | unaffected |
| 2 | GSM878293 1 | male | 30 | unaffected |
| 3 | GSM878275 1 | male | 30 | unaffected |
| 4 | GSM878257 1 | male | 35 | unaffected |
| 5 | GSM878252 1 | male | 35 | unaffected |
| 6 | GSM878239 1 | male | 35 | unaffected |
| 7 | GSM878261 1 | male | 40 | unaffected |
| 8 | GSM878258 1 | male | 45 | unaffected |
| 9 | GSM878221 1 | male | 45 | unaffected |
| 10 | GSM878301 1 | male | 50 | unaffected |
| 11 | GSM878288 1 | male | 50 | unaffected |
| 12 | GSM878273 1 | male | 50 | unaffected |
| 13 | GSM878240 1 | male | 50 | unaffected |
| 14 | GSM878230 1 | male | 50 | unaffected |
| 15 | GSM878242 1 | male | 55 | unaffected |
| 16 | GSM878236 1 | male | 55 | unaffected |
| 17 | GSM878336 1 | male | 60 | unaffected |
| 18 | GSM878322 1 | male | 60 | unaffected |

| 2B - Unaffected (female) | | | | |
|---|---|---|---|---|
| No | Sample ID | Sex | Age | Diagnosis |
| 1 | GSM878354 1 | female | 30 | unaffected |
| 2 | GSM878353 1 | female | 30 | unaffected |
| 3 | GSM878314 1 | female | 35 | unaffected |
| 4 | GSM878313 1 | female | 35 | unaffected |
| 5 | GSM878286 1 | female | 35 | unaffected |
| 6 | GSM878260 1 | female | 35 | unaffected |
| 7 | GSM878307 1 | female | 40 | unaffected |
| 8 | GSM878245 1 | female | 40 | unaffected |
| 9 | GSM878347 1 | female | 45 | unaffected |
| 10 | GSM878263 1 | female | 45 | unaffected |
| 11 | GSM878226 1 | female | 45 | unaffected |
| 12 | GSM878281 1 | female | 50 | unaffected |
| 13 | GSM878335 1 | female | 55 | unaffected |
| 14 | GSM878334 1 | female | 55 | unaffected |
| 15 | GSM878333 1 | female | 55 | unaffected |
| 16 | GSM878332 1 | female | 55 | unaffected |
| 17 | GSM878331 1 | female | 55 | unaffected |
| 18 | GSM878330 1 | female | 55 | unaffected |
| 19 | GSM878345 1 | female | 70 | unaffected |

Images 2A – 2B. Unaffected Subject Subgroups, matched by age and sex.

| 2C - Bipolar (male) | | | | |
|---|---|---|---|---|
| No | Sample ID | Sex | Age | Diagnosis |
| 1 | GSM878311 1 | male | 30 | bipolar |
| 2 | GSM878231 1 | male | 30 | bipolar |
| 3 | GSM878214 1 | male | 30 | bipolar |
| 4 | GSM878356 1 | male | 35 | bipolar |
| 5 | GSM878272 1 | male | 35 | bipolar |
| 6 | GSM878229 1 | male | 35 | bipolar |
| 7 | GSM878227 1 | male | 40 | bipolar |
| 8 | GSM878289 1 | male | 45 | bipolar |
| 9 | GSM878248 1 | male | 45 | bipolar |
| 10 | GSM878329 1 | male | 50 | bipolar |
| 11 | GSM878326 1 | male | 50 | bipolar |
| 12 | GSM878264 1 | male | 50 | bipolar |
| 13 | GSM878220 1 | male | 50 | bipolar |
| 14 | GSM878342 1 | male | 55 | bipolar |
| 15 | GSM878233 1 | male | 55 | bipolar |
| 16 | GSM878255 1 | male | 60 | bipolar |
| 17 | GSM878251 1 | male | 65 | bipolar |

| 2D - Bipolar (female) | | | | |
|---|---|---|---|---|
| No | Sample ID | Sex | Age | Diagnosis |
| 1 | GSM878344 1 | female | 25 | bipolar |
| 2 | GSM878267 1 | female | 30 | bipolar |
| 3 | GSM878284 1 | female | 35 | bipolar |
| 4 | GSM878235 1 | female | 35 | bipolar |
| 5 | GSM878243 1 | female | 40 | bipolar |
| 6 | GSM878298 1 | female | 45 | bipolar |
| 7 | GSM878271 1 | female | 45 | bipolar |
| 8 | GSM878269 1 | female | 45 | bipolar |
| 9 | GSM878339 1 | female | 50 | bipolar |
| 10 | GSM878282 1 | female | 50 | bipolar |
| 11 | GSM878278 1 | female | 50 | bipolar |
| 12 | GSM878225 1 | female | 50 | bipolar |
| 13 | GSM878270 1 | female | 55 | bipolar |
| 14 | GSM878325 1 | female | 60 | bipolar |
| 15 | GSM878244 1 | female | 60 | bipolar |
| 16 | GSM878232 1 | female | 60 | bipolar |
| 17 | GSM878266 1 | female | 65 | bipolar |
| 18 | GSM878265 1 | female | 65 | bipolar |

Images 2A – 2B. Bipolar Disorder patient Subgroups, matched by age and sex.

| Metric | Mean estimate | 95% confidence interval | Unadjusted estimate | Base line | Statistically significant |
|---|---|---|---|---|---|
| Accuracy | **0.843** | [ 0.763, 0.894 ] | 0.861 | 0.486 | ✓ |
| Balanced Accuracy | **0.846** | [ 0.765, 0.903 ] | 0.864 | 0.500 | ✓ |
| F1 Score | **0.852** | [ 0.773, 0.912 ] | 0.760 | NaN | – |
| Matthews correlation | **0.619** | [ 0.406, 0.774 ] | 0.641 | 0.000 | ✓ |
| Precision | **0.906** | [ 0.834, 0.969 ] | 0.921 | NaN | – |
| True Positive Rate | **0.691** | [ 0.452, 0.834 ] | 0.674 | 0.000 | ✓ |
| Specificity | **0.921** | [ 0.850, 0.979 ] | 0.931 | 1.000 | ✓ |
| True Positives (Out-of-sample TP rate) | **0.334** | [ 0.219, 0.419 ] | 0.325 | 0.000 | ✓ |
| True Negatives (Out-of-sample TN rate) | **0.474** | [ 0.412, 0.538 ] | 0.478 | 0.514 | – |
| False Positives (Out-of-sample FP rate) | **0.041** | [ 0.011, 0.075 ] | 0.036 | 0.000 | ✓ |
| False Negatives (Out-of-sample FN rate) | **0.152** | [ 0.086, 0.270 ] | 0.161 | 0.486 | ✓ |
| Average F1 Score | **0.873** | [ 0.802, 0.928 ] | 0.804 | NaN | – |

Image 3. Threshold Dependent metrics of the analysis.

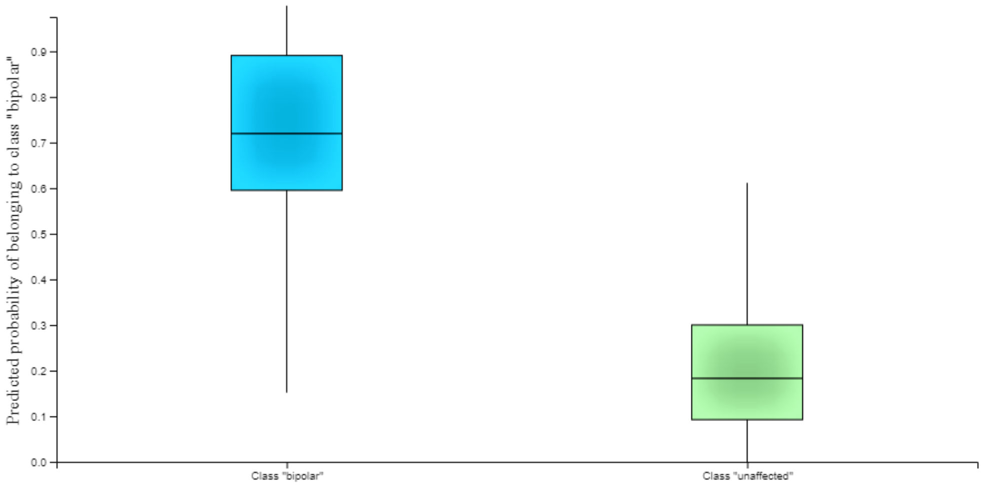


Image 4. Box Plot of the analysis.

**APPENDIX 1**

**CNS functions for the genes RNU6-576P , MIR194-2 / MiR-194-5p , GDPD5**

**1 . RNU6-576P in the CNS**

(Small nuclear RNA, pseudogene)

1. Pathological Epilepsy Overexpression Cortex (mesial temporal lobe) Hippocampus [1]
2. Non-Pathological Development - 5 five postnatal years (mainly) Dorsolateral Prefrontal Cortex hypermethylation (neurons) hypomethylation (Glia) [2]
3. Possible connection to autism [3]
4. Contribution to Psychiatric-Immune Genetic Correlation (schizophrenia – Crohn’s Disease) [4]

**2. MIR194-2 / Hsa-MiR-194-5p / MiR-194-5p in the CNS ***
(micro RNA)

1. Schizophrenia- related (in mice disease model) Downregulation Prefrontal Cortex Hippocampus [5] , [6]
2. Major Depression, Important classification feature (Patients vs Controls), Peripheral Blood [7]
3. Stress- induced Depression , differential expression (in mice disease vs control models) amygdale [8]
4. Alzheimers Disease , cmiRNA , downregulation , Cerebrospinal fluid [9]
5. Epilepsy , downregulation , serum [10] , [11]
6. Temporal lobe epilepsy , downregulation, serum [10] , [12]
7. Epilepsy (rat model) , downregulation , hippocampus [10] , [13]
8. Focal and generalized epilepsy, downregulation, serum [14] , [15]
9. Treatment Resistant Epilepsy (vs treatment responding epilepsy and healthy controls) , downregulation, serum/plasma [16] , [17]
10. Temporal lobe epilepsy (children), downregulation, peripheral blood [18]
11. Temporal lobe epilepsy (rat model), regulation of the proliferation and apoptosis of neurons in the hippocampus , neuronal cell culture [18].
12. Focal cortical dysplasia (epilepsy one of the core symptoms) , dowregulation , serum exosomes [19]
13. 22q11 Deletion Syndrome , downregulation , peripheral blood leukocytes [20]
14. lipopolysaccharide (LPS)-induced astrocytes , downregulation , cell culture  [21]
15. Autism Spectrum Disorder , Upregulation , lymphoblastoid cell lines [22] , [23]

**3. GDPD5 in the CNS**

1. Treatment Responsive Schizophrenia , Upregulation , Blood Plasma , Unknown Mechanism [24]
2. Schizophrenia , Hypomethylation , Superior Frontal Gyrus , Tissue , DNA methylation [25]
3. Epilepsy / Status Epilepticus (mouse model) , Upregulation, Hippocampus , Post-mortem , RNA expression [26]
4. Tissue samples (healthy subjects), healthy gene expression identification, human brain, hydrolysis of deacylated glycerophospholipids to glycerol phosphate and alcohol, overexpression leads to suppression of SRE-mediated transcriptional activation/ possible negative regulation role in MAPK signaling pathway.

* Hsa-MiR-194-5p is an alias of both Mir-194-1 and Mir-194-2.

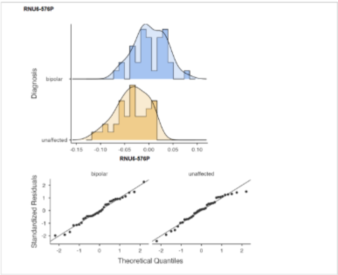


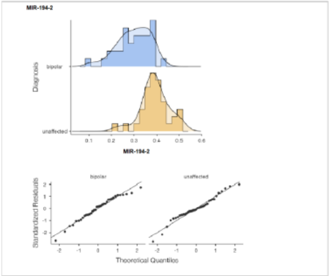


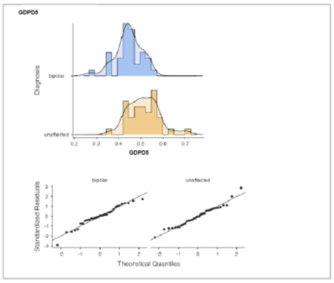

Image 5. Welsh t- test for RNU-576P, MIR194-2, GDPD5 expression differences between BD patients and Unaffected Controls.

### SI-Appendix 2

#### JADBio Description of Performed Analysis

Visit analysis

##### Setup

JADBio version **1.4.14** ran on dataset **GSE35978_BDHC_CEREBELLUM_homog_nocontrols** with **72** samples and **28869** features to create a predictive model for outcome named **Diagnosis**. The outcome was discrete leading to a **classification** modeling.

The preferences of the analysis were set to **true** for feature selection and **false** for full feature models tried.
The **AUC** metric was used to optimize for the best model.
The maximum number of features to select was set to **25**.
The effort to spend on tuning the algorithms were set to **Extensive**.
The number of CPU cores to use for the analysis was set to **1**.
The execution time was **02:16:25**.

#### Configuration Space

JADBio's AI decide to try the following algorithms and tuning hyper-parameter values:

| Algorithm Type | Algorithm | Hyper-parameter | Set of Values |
|---|---|---|---|
| Preprocessing | Contant Removal | | |
| | Standardization | | |
| Feature Selection | Test-Budgeted Statistically Equivalent Signature (SES) | alpha | 0.05, 0.1, 0.01 |
| | | maxk | 3, 2 |
| | LASSO | penalties | 1.25, 2.0, 0.0, 0.25, 1.0, 1.5, 0.5 |
| Modeling | Linear Support Vector Machines | costs | 0.1, 1.0, 0.001, 100.0, 10.0, 0.01 |
| | Polynomial Support Vector Machines | gammas | 1.0, 0.001, 0.1, 0.01, 10.0, 100.0 |
| | | costs | 0.1, 1.0, 0.001, 100.0, 10.0, 0.01 |
| | | degrees | 4, 2, 3 |

| Algorithm Type | Algorithm | Hyper-parameter | Set of Values |
|---|---|---|---|
| | RBF Support Vector Machines | gammas | 1.0, 0.001, 0.1, 0.01, 10.0, 100.0 |
| | | costs | 0.1, 1.0, 0.001, 100.0, 10.0, 0.01 |
| | Logistic Regression | lambdas | 1.0, 0.001, 0.01, 100.0, 10.0, 0.1, 1.0E-4 |
| | Random Forests | min leaf sizes | 1, 4, 3, 5, 2 |
| | | vars to split | 0.816 sqrt ( nvars ), 1.0 sqrt ( nvars ), 1.291 sqrt ( nvars ), 0.577 sqrt ( nvars ), [illegible] sqrt ( nvars ), 1.154 sqrt ( nvars ) |
| | | splits to perform | 1.0 |
| | | ntrees | 1000, 100 |
| | Decision Tree | min leaf sizes | 2, 1, 4, 3, 5 |
| | | vars to split | nvars // 1.0 |
| | | splits to perform | 1.0 |
| | | alphas | 0.1, 0.05, 0.01 |

This process led to **3017** combinations and corresponding configurations (machine learning pipelines) to try.

Configuration Estimation Protocol

JADBio’s AI system decided to estimate the out-of-sample performance of the models produced by each configuration using **Repeated 10-fold CV without dropping (max. repeats = 20).** Overall, 3017 configurations × 20 repeats × 10 folds = 603400 models were set out to train. Out of those, only 211190 models were eventually trained, as JADBio stopped all configuration evaluations when it deemed that no sufficient progress was made. JADBio **did not use** the Early Dropping criterion.
